# Neuroinflammatory and immune responses in infants with enterovirus and parechovirus meningitis, and their association with CSF pleocytosis

**DOI:** 10.1101/2025.10.31.25339167

**Authors:** Clare Mills, Holly Drummond, Helen Groves, Etimbuk Umana, Tom Waterfield

## Abstract

**Objectives:** Enterovirus (EV) and parechovirus (PeV) meningitis are significant causes of illness in febrile infants. The clinical significance of detecting EV/PeV in the CSF, particularly in the absence of pleocytosis and neurological symptoms, remains uncertain in this age group. We aimed to characterise the relationship between clinical features, systemic neuroinflammatory and immune responses, and CSF pleocytosis in infants with EV/PeV meningitis.

**Methods:** Prospective multicentre observational cohort study of febrile infants <90 days old undergoing CSF testing for infection. Infants were recruited as part of the Febrile Infant Diagnostic Assessment and Outcome (FIDO) Study from 35 Paediatric Emergency Research in the UK and Ireland (PERUKI) sites from 6th July 2022 to 31st August 2023 (<u>NCT05259683</u>). Plasma proteomic profiling of 724 inflammation and neurology-related proteins was performed using Olink technology and compared with clinical and laboratory data.

**Results:** 603 febrile infants were included, with 21/603 (3·5%) PeV, and 173/197 (28.7%) EV meningitis cases. Lymphopenia was significantly more common in infants without pleocytosis, 62/101 (63.9%), than those with pleocytosis 7/54 (16.7%). No significant differences in clinical symptoms or timing of presentation were observed with differing CSF WBC counts. Proteomic analysis (n=131) revealed that infants without pleocytosis exhibited elevated inflammatory cytokine responses and enrichment of pathways related to apoptosis and CNS involvement. In contrast, those with pleocytosis showed muted cytokine responses.

**Conclusions:** The absence of CSF pleocytosis in EV and PeV meningitis may result from systemic lymphocyte depletion, potentially mediated by cytokine-driven apoptosis. Our findings do not support the hypothesis that the lack of pleocytosis is primarily attributable to early presentation or to incidental detection of viral RNA due to viremia in infants.

## Introduction

Enterovirus (EV) and parechovirus (PeV) meningitis are increasingly recognised as important causes of febrile illness in young infants, especially as diagnostic testing expands due to the low risk of invasive bacterial infection (IBI) in those with confirmed viral meningitis ^1-4^. While outcomes are generally favourable, there remains a low but clinically significant risk of severe illness in this age group, and evidence on more subtle long-term outcomes is mixed ^5-8^. Viral meningitis is typically defined as the detection of a virus in the cerebrospinal fluid (CSF), despite many infants lacking CSF pleocytosis and neurological symptoms, compared to older children ^2,6,9^.

Viral meningitis without CSF pleocytosis is often attributed to the short interval between symptom onset and clinical presentation in infants, whereby proinflammatory cytokines have not yet attracted white blood cells (WBCs) into the CSF ^6,10^. However, studies relating symptom duration to CSF pleocytosis are mixed ^9,11-14^. Infants with viral meningitis and CSF pleocytosis absence consistently exhibit lower peripheral WBC counts compared to those with pleocytosis. This has also been attributed to early presentation time, as well as immune system immaturity in infants, whereby a robust inflammatory response is not generated due to young age ^6,9,12,15^. It is also feasible that both lower peripheral WBC counts and lack of CSF pleocytosis could be due to a virus-induced suppression of WBCs, which can occur through several mechanisms ^2,16^.

Alternatively, studies assessing viral load and genotype in children with viral meningitis have indicated that there may be two distinct infection profiles, related to virus type. Namely, infections with CNS involvement or primarily systemic infections where viral RNA may have leaked into the CSF and is a coincidental finding of viremia ^14,17,18^. Indeed, there remains no consensus on the mechanisms underlying the absence of pleocytosis in infants, nor on whether the sole detection of EV or PeV RNA in the CSF reliably indicates a true viral neurological disease.

This study aimed to characterise the relationship between systemic neuroinflammatory and immune responses, CSF pleocytosis, and symptom duration in infants with EV and PeV meningitis. By defining these associations, we sought to clarify the mechanistic basis for the absence of CSF pleocytosis and to inform the clinical interpretation and management of viral meningitis in early infancy.

## Methods

### Study design

Study participants for this analysis are from the Febrile Infant Diagnostic Assessment and Outcome (FIDO) Study. The FIDO study was a multicentre, prospective, observational cohort study that enrolled infants aged 90 days or younger who presented with a fever ≥38°C to 35 paediatric emergency departments (EDs) between July 2022 and August 2023, within the UK and Ireland. Details of the study’s protocol, methodology, recruitment process, and results have been published previously and adhere to the STROBE guidelines^19,20^. A study overview and analysis workflow are shown in Supplemental Figure 1. The FIDO study was registered at https://www.clinicaltrials.gov (trial registration: NCT05259683) on the 28th February 2022. Study ethical approval was obtained from the Office for Research Ethics Committees Northern Ireland Health and Social Care Research Ethics Committee B (Ref: 22/NI/0002), Public Benefit and Privacy Panel for Health and Social Care Scotland (Ref: 2122-0257) and Children’s Health Ireland Research and Ethics Committee Ireland (Ref: REC-082-22).

### Participants

FIDO participants were included in this study if they underwent a lumbar puncture and complete CSF testing (bacterial culture or PCR and viral PCR) at the local hospital site as part of routine care (Supplemental Figure 1). Participants were included in the Olink cohort if they had a sufficient volume of plasma sample available. All participants gave written informed consent for the storage and use of plasma samples as per the study consent procedures ^19,20^. Participants with evidence of bacterial infection (IBI or urinary tract infection) were excluded from the Olink cohort as they would have contributed significantly to plasma proteome responses in the virus-negative CSF group (Supplemental Figure 1). IBI and urinary tract infection were defined as per the FIDO study protocol ^20^.

### Procedures

Study data collection methods and Case Report Forms (CRFs) for the FIDO study have been described previously ^19,20^. An initial CRF was prospectively completed to record data regarding clinical features at the first clinician assessment by the treating clinicians. Signs of meningism were defined as any of the following: bulging fontanelle, neck stiffness, seizures or focal neurological signs. Unwell appearance was defined as abnormal global assessment and/or abnormal vital signs (based on APLS recommendations for age). Fever duration was recorded as the number of hours from first fever in categories of 0-6, 6-12 and 12-24 and >24 hours.

A follow-up CRF was completed seven days after discharge to record laboratory results and length of stay. All laboratory testing (culture, PCR, WBC counts, and CRP measurements) was conducted at local hospital laboratories using commercially available, routinely used tests on samples collected at initial presentation, as part of routine care. CSF pleocytosis was defined as >15 WBCs per mm3 in the CSF. If > 500/mm3 red blood cells were recorded in the CSF a ratio of 500:1 white to red blood cells was applied. Lymphopenia was defined as a peripheral lymphocyte count lower than age-based reference ranges (0-28 days; 2·8-9.1x10^9^/l, 29-90 days; 4·0-10·0x10^9^/l).

EDTA plasma samples were collected during routine phlebotomy on initial presentation to ED and stored at -80C, where possible. Plasma concentrations of 768 proteins were measured using the Olink® platform (Uppsala, Sweden) “Explore Neurology” and “Inflammation” antibody-based Proximity Extension Assays. Analysis was performed by an Olink Certified Service Provider (Randox Laboratories, Antrim, UK). Data were provided normalised and log2 transformed before analysis. Proteins were removed if they were detected in less than 50% of samples, and duplicate proteins were removed from the Neurology panel. 724 proteins were included for final analysis.

### Statistical analysis of clinical data

Categorical clinical variables were reported as frequencies and percentages, with differences among groups analysed by Fisher’s exact test. Continuous variables were reported as mean and standard deviation (SD) for normally distributed data (based on the Shapiro–Wilk normality test), or median and interquartile ranges (IQR) for non-normally distributed data. Missing data were excluded. Differences among groups were analysed using Student’s t-test or the Mann–Whitney U-test, depending on the normality of the data. Significance was based on a false discovery rate (FDR) using Benjamini–Hochberg correction of less than 0.05. Analysis was performed using GraphPad Prism®version 10.0.0 for Windows (GraphPadSoftware, Boston, MA, USA).

### Statistical analysis of Olink data

Olink data analysis was conducted in R (v4.3.3). Differentially expressed proteins between infection groups were identified using the R package “Olink^®^ Analyze”. Significance was based on a FDR using Benjamini–Hochberg correction of less than 0.05. Pearson correlation coefficients between proteins and indicated cell counts were calculated using the R function “cor”. p-values from correlation analysis were adjusted based on an FDR (0.05) using the “p.adjust” function (Benjamini–Hochberg correction). To generate hierarchically clustered heatmaps, the data were first scaled per protein using “scale”. Hierarchically clustered heatmaps were then generated using “ComplexHeatmap” with the HeatmapAnnotation() function to visualise corresponding clinical data.

For functional enrichment analysis differentially expressed proteins were input into the Metascape Gene Annotation and Analysis Resource (http://metascape.org) to query Kyoto Encyclopedia of Genes and Genomes (KEGG), Gene Ontology (GO) and Reactome Gene Sets resources. The following criteria were used for enrichment analysis; minimum count of 3, enrichment factor of >1.5, p< 0.01. Terms significantly enriched in the indicated protein sets were automatically clustered by Metascape into nonredundant groups and presented as bar charts ^21^.

## Results

### Patient characteristics and clinical comparisons

603 infants were included in this study, with 197/603 (32.5%) cases of virus-positive CSF, with a prevalence of 21/603 (3.5%) PeV, and 173/603 (28.7%) EV (Supplemental Figure 1). 87/603 (14%) infants underwent viral blood PCR testing. Patient demographics and clinical features are summarised in Table 1. No significant differences in age or sex were observed between virus-positive and virus-negative groups, or between EV and PeV infections. IBI was more frequent in virus-negative CSF infants (35/406, 8.5%) compared to virus-positive (1/197, 0.5%, P = 0.003), with only one IBI case in an infant with EV-positive CSF. Compared to virus-negative infants, those with EV or PeV-positive CSF exhibited increased incidence of rash (P =0.003) and fewer respiratory symptoms (P = 0.003). 11/603 (1.8%) infants in the full cohort with complete CSF testing had signs of meningism.

**Table 1.** Patient characteristics and demographics. *Statistically significant; FDR<0.05, **Viruses in CSF: EV n=173, PeV n=21, Adenovirus n=1, SARS-CoV-2 n=1, Human herpesvirus 6 n=2. *CRP, C-reactive protein; CSF, cerebrospinal fluid; EV, Enterovirus; GI, gastrointestinal; IBI, invasive bacterial infection; IQR, interquartile range; LOS, length of stay; PeV, Parechovirus; WBC, white blood count*.

|  | Complete CSF testing | Virus positive CSF** | Virus negative CSF | Adjusted P Value (CSF virus + vs -) | PeV positive CSF | EV positive CSF | Adjusted P Value (PeV vs EV CSF) |
| --- | --- | --- | --- | --- | --- | --- | --- |
| **Total, n (%)** | 603 | 197 (32.7) | 406 (67.3) |  | 21 (3.5) | 173 (28.7) |  |
| **Males, n (%)** | 365 (60.5) | 117 (59.4) | 248 (61.1) | 0.812 | 12 (57.1) | 104 (60.1) | 0.919 |
| **Age (days) Median (IQR)** | 39 (24-58) | 36 (25-53) | 41 (24-59) | 0.102 | 29 (17-49) | 36 (25-53) | 0.295 |
| **Temperature(°C) Median (IQR)** | 38.1 (37.6-38.5) | 38.2 (37.8-38.5) | 38.0 (37.5-38.5) | 0.073 | 38.0 (37.6-38.6) | 38.2 (37.9-38.5) | 0.905 |
| **Unwell Appearance n (%)** | 405 (67.2) | 140 (71.1) | 265 (65.3) | 0.213 | 16 (76.2) | 124 (71.7) | 0.919 |
| **Signs of Meningism n (%)** | 11 (1.8) | 3 (1.5) | 8 (2.0) | 0.999 | 0 (0.0) | 3 (1.7) | 0.999 |
| **Rash n (%)** | 120 (19.9) | 56 (28.4) | 64 (15.8) | 0.003* | 5 (23.8) | 51 (29.5) | 0.919 |
| **GI Symptoms n (%)** | 176 (29.2) | 46 (23.4) | 130 (32.0) | 0.056 | 4 (19.0) | 42 (24.3) | 0.919 |
| **Behaviour Change n (%)** | 473 (78.4) | 164 (83.2) | 309 (76.1) | 0.075 | 20 (95.2) | 141 (81.5) | 0.295 |
| **Respiratory Symptoms, n (%)** | 215 (35.7) | 41 (20.8) | 174 (42.9) | 0.003* | 3 (14.3) | 38 (22.0) | 0.904 |
| **CSF WBC count (x10<sup>9</sup>/L) Median (IQR)** | n=548<br>2.0 (1.0-7.0) | n=177<br>4.0 (1.0-46.5) | n= 371<br>1.0 (1.0-5.0) | 0.003* | n=19<br>1.0 (1.0-5.0) | n=155<br>5.0 (1.0-82.0) | 0.105 |
| **CSF pleocytosis n (%)** | 76 (12.6) | 55 (31.1) | 21 (5.9) | 0.003* | 0 (0.0) | 54 (34.8) | 0.005* |
| **Peripheral WBC count (x10<sup>9</sup>/L) Median (IQR)** | n=578<br>9.6 (6.8-12.9) | n=189<br>8.6 (6.6-11.4) | n=389<br>10.3 (7.1-13.9) | 0.003* | n=19<br>5.7 (4.8-8.1) | n=167<br>8.8 (6.8-11.6) | 0.005* |
| **Lymphocyte count (x 10<sup>9</sup>/L) Median (IQR)** | n=577<br>3.7 (2.4-5.4) | n=189<br>3.1(2.0-4.7) | n=388<br>4.0(2.5-5.7) | 0.003* | n=19<br>1.9 (1.2-3.2) | n=167<br>3.2 (2.1-4.7) | 0.018* |
| **Lymphopenic n (%)** | 236 (39.1) | 91 (48.1) | 146 (37.6) | 0.043* | 15 (79.0) | 75 (44.9) | 0.005* |
| **Neutrophil count (x 10<sup>9</sup>/L) Median (IQR)** | n=577<br>4.0 (2.5-6.6) | n=189<br>4.0 (2.8-5.9) | n=388<br>3.91 (2.4-7.2) | 0.889 | n=19<br>2.8 (1.7-3.4) | n=167<br>4.19 (3.0-6.0) | 0.005* |
| **CRP (mg/L) Median (IQR)** | n=595<br>10.0 (4.0-29.0) | n=195<br>10.0 (4.0-22.0) | n=400<br>10.7 (3.0-44.8) | 0.209 | n=21<br>5.0 (4.0-10.0) | n=171<br>10.0 (5.0-22.0) | 0.018* |
| **LOS (days) Median (IQR)** | 3 (2-4) | 3 (2-4) | 3 (2-4) | 0.288 | 3 (3-4) | 3 (2-4) | 0.295 |
| **IBI n (%)** | 36 (6.0) | 1 (0.5) | 35 (8.6) | 0.003* | 0 (0.0) | 1 (0.6) | 0.999 |

Peripheral WBC counts were lower in infants with virus-positive CSF than in infants with virus-negative CSF (8.6 vs 10.3x10^9^/L, P=0.003), and lower in PeV than EV infections (5.7 vs 8.8x10^9^/L, P=0.008). The difference was attributable to reduced peripheral lymphocytes in virus-positive vs virus-negative groups (3.1 vs 4.0 ×10^9^/L, P = 0.003), with neutrophil counts comparable between these two groups (P = 0.845). Lymphopenia was more prevalent in virus-positive vs virus-negative infants (48.1% vs 37.6%, P = 0.043) and in PeV vs EV infections (79.0% vs 44.9%, P = 0.005). CSF pleocytosis occurred in 54/155 (34.8%) EV-positive infants but was absent in all PeV-positive cases (0/19, 0%). Clinical and laboratory features grouped by pleocytosis status in EV-positive infants are detailed in Table 2. Infants without pleocytosis had reduced lymphocytes (2.7 vs 4.7 ×10^9^/L, P = 0.004), and a higher prevalence of lymphopenia (63.9% vs 16.7%, P=0.004). No other significant clinical symptom differences were noted between groups.

**Table 2.** Patient characteristics of infants with EV detected in the CSF, with and without CSF pleocytosis. *Statistically significant; FDR<0.05. *IQR, interquartile range; GI, gastrointestinal; CSF, cerebrospinal fluid; WBC, white blood cell; CRP, C-reactive protein; LOS, length of stay; IBI, EV, Enterovirus*.

|  | EV without CSF<br>pleocytosis | EV with CSF<br>pleocytosis | Adjusted P<br>value |
| --- | --- | --- | --- |
| **Total n (%)** | 101 (65.2) | 54 (34.8) |  |
| **CSF WBC count (x10<sup>9</sup>/L)**<br>Median (IQR) | n=101<br>1.0 (1.0-4.0) | n=54<br>210 (72.8-403.0) |  |
| **Males n (%)** | 60 (59.4) | 33 (61.1) | 0.999 |
| **Age (days) Median (IQR)** | 36 (25-56) | 38 (24-53) | 0.999 |
| **Temperature (°C) Median (IQR)** | 38.2 (37.9-38.5) | 38.2 (37.8-38.5) | 0.999 |
| **Unwell Appearance n (%)** | 73 (72.3) | 38 (70.4) | 0.999 |
| **Signs of Meningism n (%)** | 1 (1.0) | 2 (3.7) | 0.675 |
| **Rash n (%)** | 30 (29.7) | 14 (25.9) | 0.999 |
| **GI Symptoms n (%)** | 21 (20.8) | 17 (31.5) | 0.485 |
| **Change in Behaviour n (%)** | 86 (85.1) | 40 (74.1) | 0.438 |
| **Respiratory symptoms n (%)** | 24 (23.8) | 10 (18.5) | 0.999 |
| **Peripheral WBC count (x10<sup>9</sup>/L)**<br>Median (IQR) | n=97<br>7.9 (6.0-10.2) | n=54<br>10.6 (8.2-12.2) | 0.004* |
| **Lymphocyte count (x 10<sup>9</sup>/L)**<br>Median (IQR) | n=97<br>2.7 (1.8-3.8) | n=54<br>4.7(3.6-6) | 0.004* |
| **Lymphopenic n (%)** | 62 (63.9) | 7 (16.7) | 0.004* |
| **Neutrophil count (x 10<sup>9</sup>/L)**<br>Median (IQR) | n=97<br>4.0 (2.7-6.2) | n=54<br>4.2 (3.1-5.5) | 0.999 |
| **CRP (mg/L)**<br>Median (IQR) | n=101<br>17.0 (8.7-28.0) | n=53<br>5.0 (2.2-9.5) | 0.004* |
| **Hospital admission n (%)** | 101 (100.0) | 54 (100.0) | 0.999 |
| **LOS, days Median (IQR)** | 3 (2-4) | 3 (2-4) | 0.784 |
| **PICU Admission n (%)** | 0 (0.0) | 0 (0.0) | 0.999 |

### Plasma response to CSF virus type

Olink proteomic analysis was performed on 131 plasma samples to characterise the host response to viral meningitis (Supplemental Figure 1). Demographic and clinical features of the Olink cohort were comparable to the full study population in terms of age (P = 0.277) and sex (P > 0.999) (Supplemental Table 1A). CSF WBC differences also remained consistent, with higher counts in virus-positive vs virus-negative infants (P < 0.001) and in EV vs PeV cases (P = 0.007). Direct comparison between infants with and without sample available for Olink analysis, within the clinical data cohort, also showed there were no significant differences between the proteomic and non-proteomic groups in key demographic or clinical characteristics, including: age, sex and clinical features at presentation (temperature, presence of rash, behavioural change, respiratory symptoms, unwell appearance) (Supplemental Table1B).

Differential expression analysis identified 32 differentially expressed proteins in PeV-positive and 11 in EV-positive CSF cases compared to virus-negative controls (Figure 1A&B, Supplemental Tables 2&3). Boxplots of significantly altered proteins are shown in Supplemental Figure 2. Figure 1C shows hierarchical clustering of the significantly differentiated plasma proteins from both the PeV-positive vs virus-negative and EV-positive vs virus-negative analysis with a fold change >1.0. Samples from PeV-positive cases clustered tightly together and displayed a uniform upregulation of the included proteins. In contrast, EV-positive samples exhibited more heterogeneous clustering patterns, with variable expression across the same protein panel. This heterogeneity appeared to align with clinical parameters, particularly pleocytosis and lymphopenia, as shown by the annotation bars. EV cases with pleocytosis tended to cluster toward lower relative expression of the differentially abundant proteins, whereas those without pleocytosis showed relatively higher expression levels. No clear clustering was observed in relation to age, sex, or fever duration.

**Figure 1.**
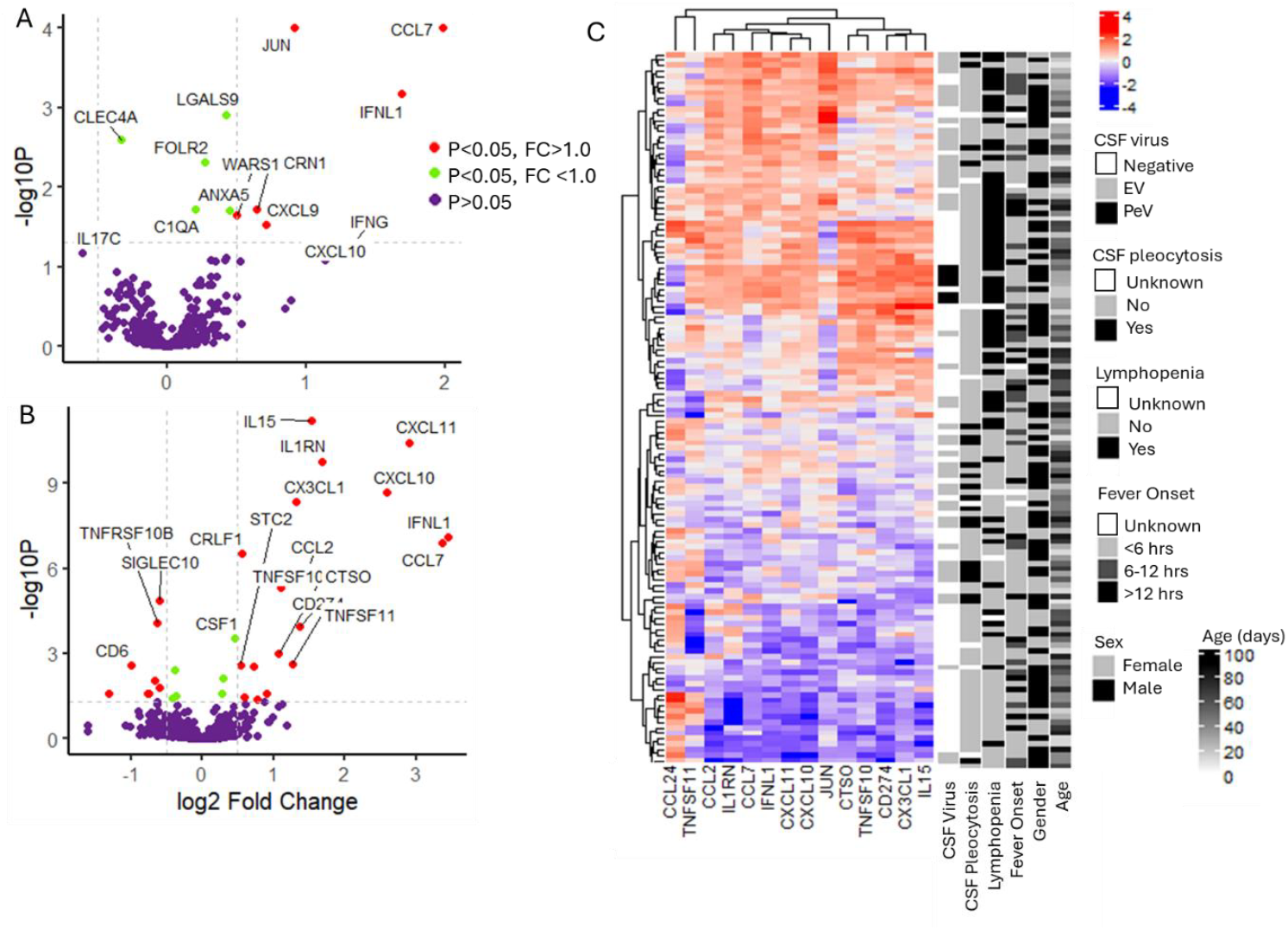
Host plasma proteome response to EV and PeV meningitis. (A&B) Volcano plots of differentially expressed proteins (FDR 0.05) between infant groups from the Olink cohort: (A) EV-positive CSF infants (n=49) and viral negative CSF infants (n=76), (B) PeV-positive CSF infants (n=6) and viral negative CSF infants (n=76). (C) Hierarchical clustering displayed as a heatmap of the Olink cohort (n=131) by significantly differentiated proteins (FDR<0.05) with a fold change of >1.0 between EV vs viral negative CSF and PeV vs viral negative CSF. Coloured by expression values. *EV; enterovirus, PeV; parechovirus, CSF; cerebrospinal fluid, FDR; false discovery rate*

### Plasma response to CSF pleocytosis and lymphopenia

As protein expression patterns appeared to relate to both CSF white cell and peripheral lymphocyte counts, and lymphopenia was the main variable distinguishing infants with and without pleocytosis, we next investigated how these parameters shaped plasma protein expression. A correlation analysis of the plasma proteome with lymphocyte and CSF WBC counts was performed in infants with virus-positive CSF. A clear plasma response was associated with both increasing and decreasing lymphocyte counts (Figure 2A) (Supplemental Table 4). Scatter graphs of correlating proteins showed a linear relationship (Supplemental Figure 3). A clear plasma response was associated with reduced CSF WBC counts, but not with increasing CSF WBC counts. 30 plasma proteins were significantly negatively correlated with CSF WBC counts (Figure 2B, Supplemental Table 5), while only three showed positive correlations. Scatter plots suggest that these positive correlations were driven by outliers rather than linear relationships (Supplemental Figure 4).

**Figure 2.**
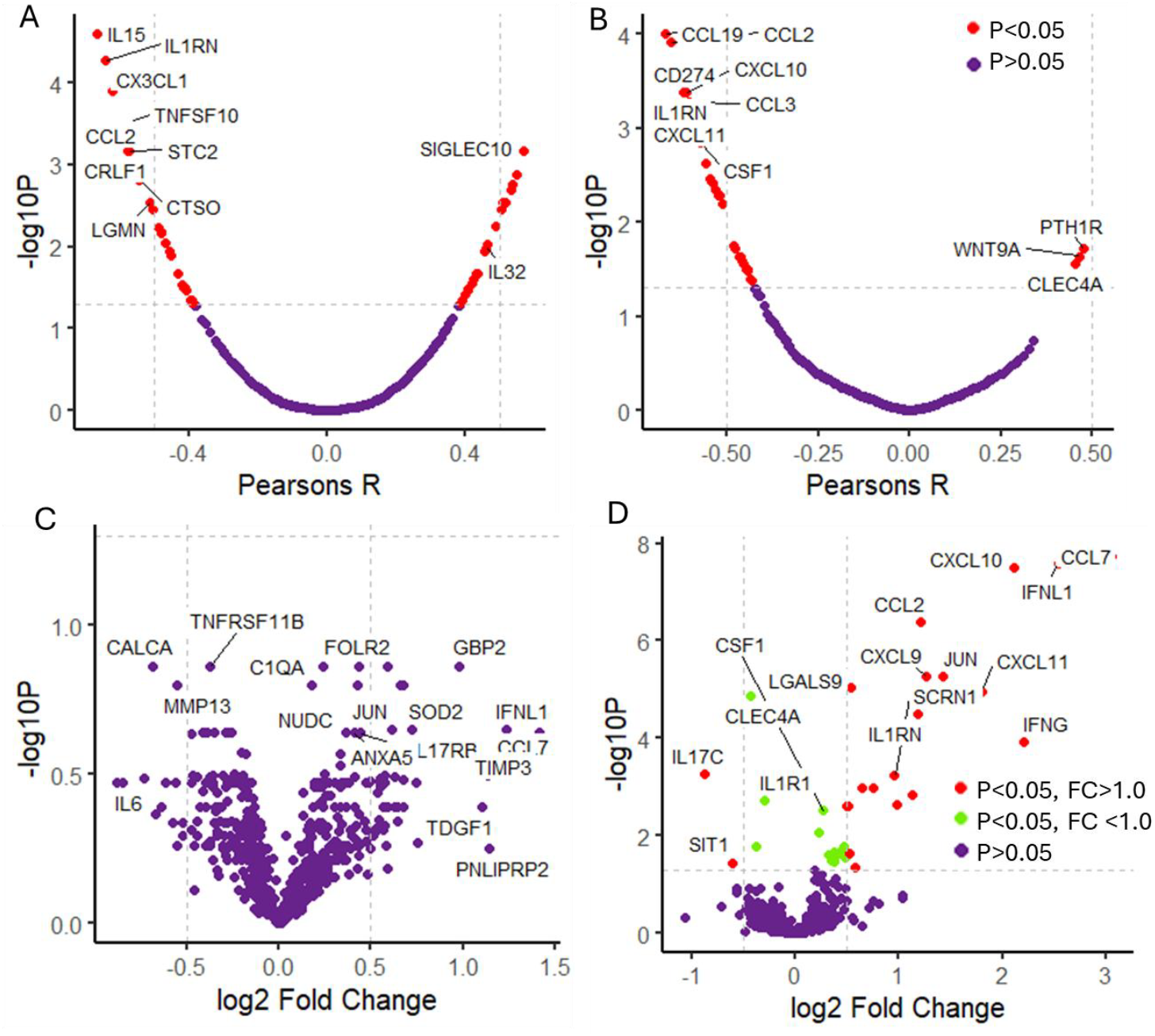
Plasma proteome response, CSF pleocytosis and peripheral lymphocyte count. (A&B) Volcano plot showing the correlation in CSF virus-positive samples between plasma protein expression and: (A) Peripheral lymphocyte count (n=54), infants without data on peripheral lymphocyte count were excluded (n=1), (B) CSF WBC count (n=49), infants without data on CSF WBCs were excluded (n=6). (C&D) Volcano plots of differentially expressed proteins (FDR 0.05) between: (C) EV-positive CSF infants with CSF pleocytosis (n=21) and virus-negative CSF (n=76), (D) EV-positive CSF infants without CSF pleocytosis (n=23) and virus-negative CSF (n=76). *EV; enterovirus, PeV; parechovirus, CSF; cerebrospinal fluid, WBC; white blood cells, FDR; false discovery rate*

To further try to identify a unique plasma response associated with CSF pleocytosis, EV-positive CSF infants were grouped by the presence or absence of CSF pleocytosis and compared to the virus-negative group. Differential expression analysis identified 35 proteins significantly altered in EV-positive infants without pleocytosis, however there were no significantly differentially expressed proteins between EV with pleocytosis and virus-negative samples with FDR <0.05 (Figure 2C&D, Supplemental Table 6). There were, however, 27 proteins identified as significant using a non-adjusted p-value < 0.01 (Supplemental Figure 5 and Table 7). 11 of the 27 proteins overlapped with the non-pleocytosis group. This suggests that FDR correction may be overly conservative in this context, masking true associations.

### Clustering of plasma response and enrichment analysis

Given the apparent influence of CSF and peripheral immune cell responses on protein expression, we next assessed how these parameters shaped plasma proteomic clustering and explored the functional enrichment of the associated protein signatures. A hierarchically clustered heatmap was used to display the expression of the 39 plasma proteins significantly correlated with either CSF WBC or peripheral lymphocyte count (FDR <0.01) (Figure 3A). The heatmap, generated with the dendrogram cut at *k = 3*, showed a clear separation of the samples into three distinct clusters of infants; Cluster 1, 2 and 3. 100% (17/17) of Cluster 1 infants had pleocytosis, in contrast to 10.6% (2/19) and 15.4% (2/13) in Clusters 2 and 3, respectively (Table 3). Cluster 2 infants were significantly younger than those in Clusters 1 and 3 (27.0 vs 41.5 and 57.5 days (P = 0.014). They also exhibited markedly lower peripheral lymphocyte counts, 1.7 × 10^9^/L, than in Clusters 1 (4.5 × 10^9^/L) and 3 (3.5 × 10^9^/L) (P = 0.004). No significant differences in sex, presenting symptoms, or hospital stay were observed among clusters.

**Figure 3.**
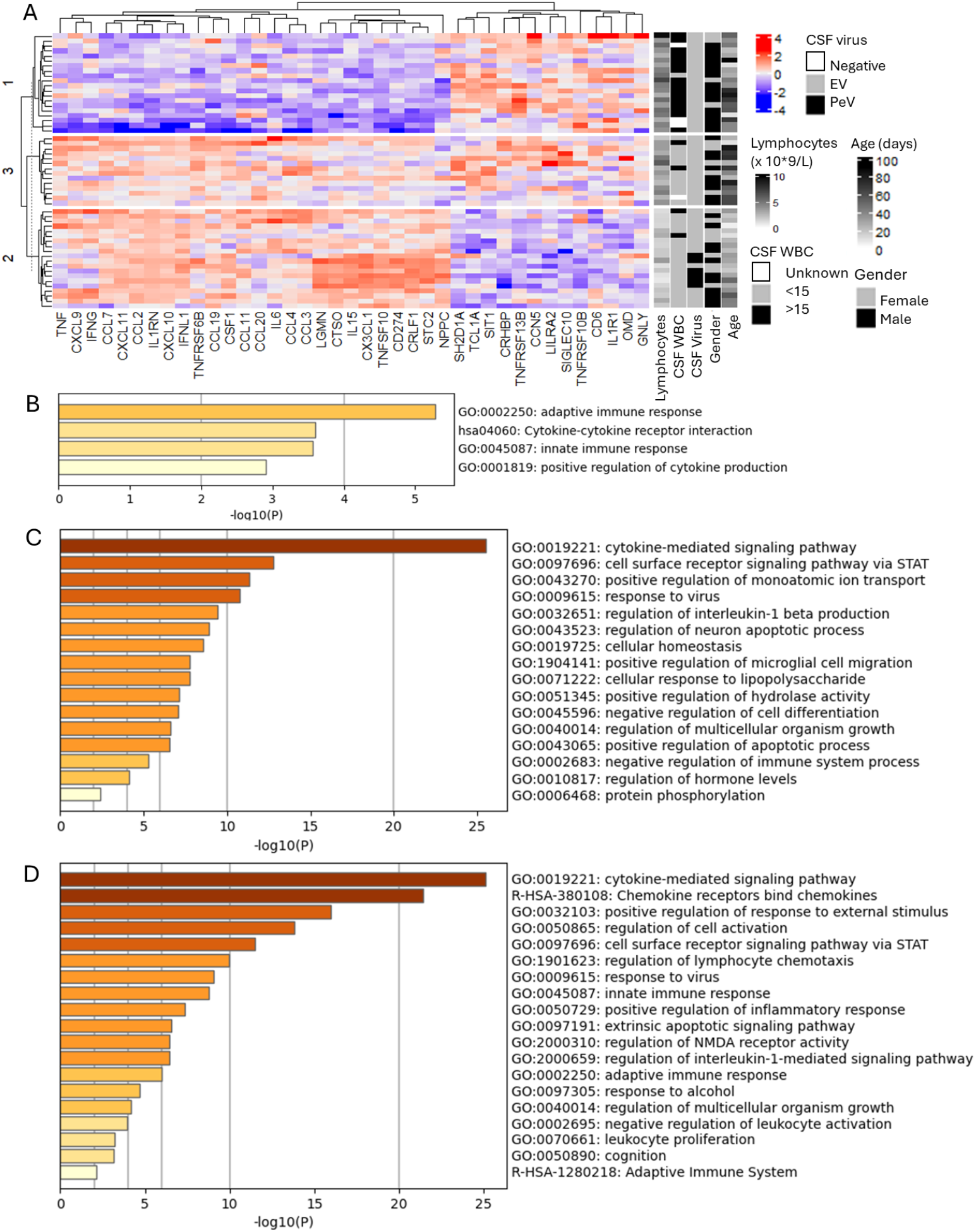
Plasma proteome clusters and functional analysis of EV/PeV meningitis. (A) Hierarchical clustering displayed as heatmap of all infants with EV or PeV positive CSF from the Olink cohort (n=55) by proteins significantly correlated with CSF WBC count or peripheral lymphocyte count, (FDR<0.05). Coloured by expression values. (B-D) Bar chart of enriched terms from proteins upregulated in heatmap cluster 1 (B), 2 (C) and 3 (D). Coloured by p-values, metascape. *EV; enterovirus, PeV; parechovirus, CSF; cerebrospinal fluid, WBC; white blood cells, FDR; false discovery rate*

**Table 3.** Patient characteristics of infants with virus positive CSF in the three clusters defined by protein expression levels, as per Figure 3B. *Statistically significant. *IQR, interquartile range; GI, gastrointestinal; CSF, cerebrospinal fluid; WBC, white blood cell; LOS, length of stay*

|  | Cluster 1 | Cluster 2 | Cluster 3 | Adjusted P value | Multiple comparisons |
| --- | --- | --- | --- | --- | --- |
| **Total n (%)** | 21 (38.2) | 20 (36.4) | 14 (25.5) |  |  |
| **Males n (%)** | 16 (76.2) | 11 (55) | 9 (64.3) | 0.404 |  |
| **Age (days) Median (IQR)** | 41.5 (28.5-66) | 27.0 (19.5-35.3) | 57.5 (25.0-70.0) | 0.014* | 1 vs 2 0.028*<br>1 vs 3 >0.999<br>2 vs 3 0.009* |
| **Temperature (°C) Median (IQR)** | 38.2 (37.8-38.5) | 38.2 (38.0-38.6) | 38.0 (37.5-38.6) | 0.741 |  |
| **Unwell Appearance n (%)** | 16 (76.2) | 17 (85.0) | 10 (71.4) | 0.762 |  |
| **Rash n (%)** | 8 (38.1) | 6 (30.0) | 5 (35.7) | 0.935 |  |
| **GI Symptoms n (%)** | 8 (38.1) | 4 (20.0) | 2 (14.3) | 0.404 |  |
| **Change in Behaviour n (%)** | 19 (90.5) | 16 (80.0) | 11 (78.6) | 0.435 |  |
| **Respiratory symptoms n (%)** | 5 (28.6) | 4 (20.0) | 5 (35.7) | 0.762 |  |
| **CSF WBC count (x10<sup>9</sup>/L) Median (IQR)** | n=17<br>257.5 (40.3-531) | n=19<br>1.0 (0.0-4.1) | n=13<br>1.0 (0.5-6.4) | 0.004* | 1 vs 2 <0.001*<br>1 vs 3 <0.001*<br>2 vs 3 >0.999 |
| **Peripheral WBC count (x10<sup>9</sup>/L) Median (IQR)** | n=20<br>10.3 (8.2-13.6) | n=20<br>6.0 (4.8-7.1) | n=14<br>8.3 (6.2-9.9) | 0.004* | 1 vs 2 <0.001*<br>1 vs 3 0.243<br>2 vs 3 0.052 |
| **Lymphocyte count (x 10<sup>9</sup>/L) Median (IQR)** | n=20<br>4.5 (3.6-5.3) | n=20<br>1.7 (1.4-2.0) | n=14<br>3.5 (3.0-4.9) | 0.004* | 1 vs 2 <0.001*<br>1 vs 3 0.397<br>2 vs 3 0.001* |
| **Lymphopenic n (%)** | 6/20 (30%) | 18/20 (90%) | 6/14 (42.9%) | 0.004* | 1 vs 2 <0.001*<br>1 vs 3 0.440<br>2 vs 3 0.003* |
| **Neutrophil count (x 10<sup>9</sup>/L) Median (IQR)** | n=20<br>4.9 (3.2-6.3) | n=20<br>3.4 (2.8-4.0) | n=14<br>3.0 (3.0-4.9) | 0.278 |  |
| **LOS, days Median (IQR)** | 3 (2-4) | 3 (2-4) | 2(1-4) | 0.356 |  |

Functional enrichment analysis of cluster-specific upregulated proteins (Supplemental Table 8) revealed distinct biological signatures. Cluster 2, despite minimal pleocytosis, was enriched for pro-inflammatory and antiviral pathways, as well as CNS-associated processes including “regulation of neuron apoptotic process” and “positive regulation of microglial cell migration” (Figure 3C). It also showed enrichment for “positive regulation of apoptotic processes.” Proteins upregulated in Cluster 1, the only group with consistent CSF pleocytosis, showed only four enriched terms, with “adaptive immune response” being the most significant (Figure 3B). Notably, no CNS-related or apoptotic pathways were enriched in this cluster.

Enriched terms in Cluster 3, contained a combination of enriched terms from both Cluster 1 and 2 (Figure 3D), with the “regulation of NMDA receptor activity” cluster representing terms relating to neuron apoptotic processes. Therefore, CNS-related and apoptotic signalling was observed only in Clusters 2 and 3, which both lacked significant pleocytosis.

### Fever duration and host response

While no association was observed between fever duration and the host proteomic response within the Olink cohort (Figure 1D), to further investigate a possible association, differential expression analysis was carried out on infants with <6 hours and >6 hours from fever onset. No differentially expressed proteins were identified between groups at FDR < 0.05 (Figure 4A), and only two proteins reached significance (p < 0.01) with an unadjusted p value. In the full clinical data cohort, CSF WBC counts did not differ significantly across fever duration categories (<6 h, 6–12 h, 12–24 h, >24 h; Figure 4B). Peripheral lymphocyte counts also showed no significant variation by fever duration (Figure 4C).

**Figure 4.**
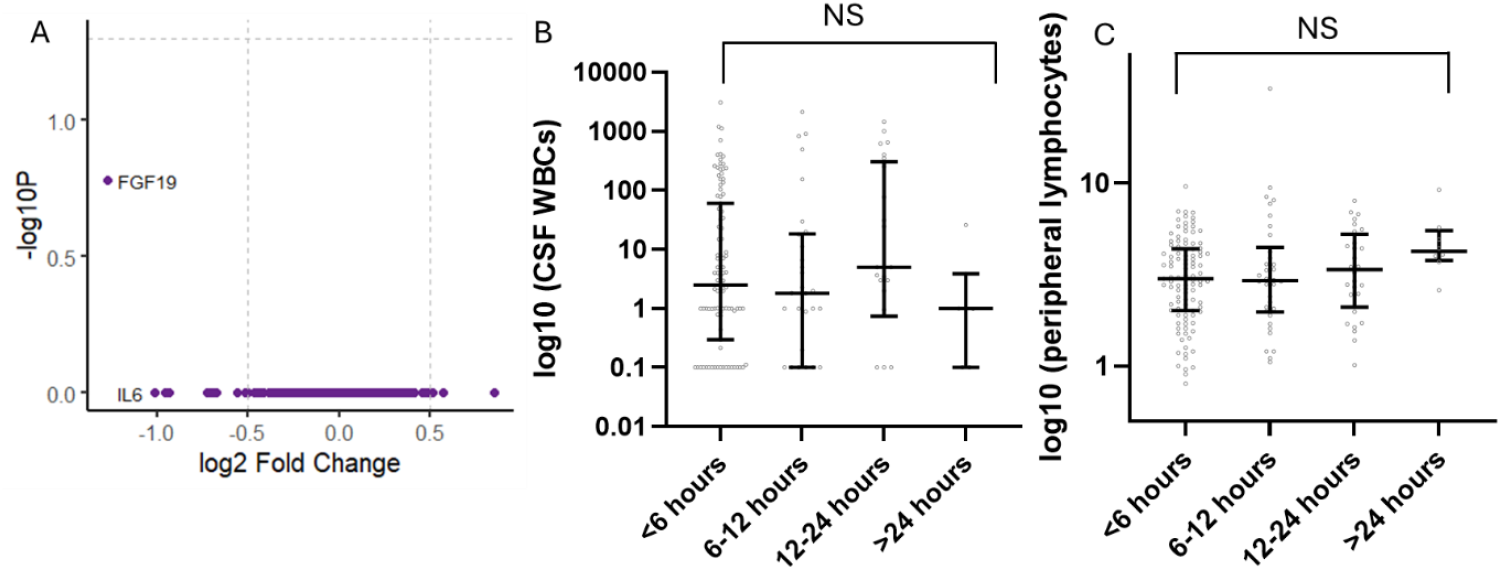
The effect of fever duration of plasma proteome response and WBC counts. (A) Volcano plot of differentially expressed proteins (FDR 0.05) between virus positive CSF infants in the Olink cohort, presenting <6 hours (n=37) vs > 6 hours (n=14) from first recorded fever. Infants without information on fever duration were excluded (n=4). (B) Box plot showing CSF WBC count by fever duration category for all participants in clinical data cohort with EV or PeV positive CSF (n=166; <6hours n=103, 6-12 hours n=30, 12-24 hours n=26, >24 hours n=7). Infants without data on fever duration or CSF WBC were excluded (n=28). (C) Box plot showing peripheral lymphocyte count by fever duration category for all participants with EV or PeV positive CSF (n=175; <6hours n=105, 6-12 hours n=34, 12-24 hours n=28, >24 hours n=8). Infants without data on fever duration or peripheral lymphocyte count were excluded (n=19). Lines represent median and interquartile ranges. *EV; enterovirus, PeV; parechovirus, CSF; cerebrospinal fluid, WBC; white blood cells, FDR; false discovery rate*

## Discussion

This study constitutes the most comprehensive characterisation to date of the host immune response and clinical characteristics of EV and PeV meningitis in infants. We have identified distinct immune response patterns in infants without CSF pleocytosis, associated with lymphopenia and unrelated to the time of disease onset. Notably, infants lacking CSF pleocytosis exhibited a pronounced systemic proinflammatory cytokine response, with plasma protein signatures overlapping pathways implicated in neuroinflammatory processes. These findings suggest that the absence of pleocytosis does not necessarily indicate a lack of CNS-directed immune activation but may instead reflect a distinct systemic immune response profile.

In our prospective multicentre cohort of febrile infants, we found a viral meningitis prevalence of 32.7%, demonstrating its significant burden. Consistent with previous studies, we highlight the difficulty in distinguishing viral meningitis from other febrile illnesses based on clinical features. Although infants with viral meningitis were statistically more likely to present with a rash, its diagnostic utility was limited as only 28.4% exhibited this feature. Classic signs of meningism were rare, indicating their limited diagnostic value in this population.

CSF pleocytosis was not observed in infants with PeV meningitis and in only 34% of those with EV meningitis, consistent with previous reports^7^. Infants without CSF pleocytosis exhibited significantly lower peripheral WBC counts, as supported by earlier studies, which was driven by a reduction in lymphocytes^6,9,12,15^. Lymphopenia was present in 79% (15/19) of PeV-infected infants, compared to 44.9% (75/167) of those with EV, indicating, along with the universal absence of CSF pleocytosis, a more consistent immunological phenotype in PeV infection.

Proteomic profiling of over 700 plasma proteins associated with inflammation and neurology mirrored these findings. PeV infection showed a uniform inflammatory signature, whereas EV responses were heterogeneous and closely linked to pleocytosis and lymphopenia. Infants with EV meningitis but without pleocytosis exhibited plasma profiles more similar to PeV infection, than EV with pleocytosis, indicating a shared mechanism underlying absent CSF pleocytosis.

Given the variation in plasma host responses relative to CSF pleocytosis and lymphopenia, these relationships were further examined. Infants with CSF pleocytosis exhibited a detectable but subdued host response, with significant protein changes emerging only after removal of FDR correction. In contrast, infants without pleocytosis showed a strong systemic cytokine activation. Clustering of plasma proteins by CSF and peripheral lymphocyte counts identified three distinct groups; clusters 1, 2 and 3.

Clusters 2 and 3, both characterised by minimal pleocytosis, demonstrated robust upregulation of cytokines and chemokines, with enrichment of inflammatory and apoptosis-related pathways. Their plasma profiles, dominated by IFNγ-inducible cytokines (CXCL9, CXCL10, CXCL11), IL-6, TNF, and IL-15, resembled a cytokine storm, an exaggerated inflammatory state linked to lymphocyte depletion and immune dysregulation after viral infection^22^. This cytokine-driven lymphopenia may limit immune cell trafficking into the CNS, explaining the absence of pleocytosis. Although enrichment terms related to neuroinflammatory and apoptotic processes were identified in these two clusters, it is worth noting that these annotations are derived from plasma protein homology and may reflect systemic pathways with overlap to CNS immune signalling rather than direct evidence of central processes. The enrichment of apoptosis-associated proteins nonetheless supports the possibility that lymphocyte apoptosis contributes to systemic lymphopenia in this context.

Clusters 2 and 3 differed in age distribution and immune pathway enrichment, suggesting that immune maturation influences the variability in host responses. Cluster 3 uniquely expressed lymphocyte-associated genes (CD6, SH2D1A, SIT1, TCL1A), and pathway enrichment for adaptive singling, indicating stronger adaptive immune engagement, whereas Cluster 2 only reflected the predominantly cytokine-driven response. This aligns with the younger age and more pronounced lymphopenia observed in Cluster 2. Cluster 1, which included most infants with CSF pleocytosis, also showed enrichment for adaptive immune signalling but lacked the cytokine-dominated inflammatory profile seen in Clusters 2 and 3.

EV and PeV meningitis are generally associated with favourable outcomes, however evidence on more subtle long-term outcomes is mixed ^5-8^. Studies of EV-A71, which can cause severe neuronal complications, show that cytokine responses correlate with disease severity, indicating that the host’s inflammatory response magnitude plays a key role in disease severity ^23^. Indeed, other severe viral infections (e.g. COVID-19) are characterised by marked depletion of lymphocytes in the blood, correlated with cytokine dysregulation ^24,25^. While this study found no indications of increased disease severity within the different clusters, we were limited to reporting length of stay as an outcome, with no long-term follow-up. Given the evidence of increased neurological disease severity with “cytokine storms” in EV-71 and other viral infections, and the increase in enriched functional terms relating to neuron apoptosis in our clusters of infants, it could be hypothesised that infants who lack CSF pleocytosis are at increased risk of worse outcomes ^22,23^. Indeed, Kadambari et al reported six infants with death or sequelae following EV meningitis, over half of whom lacked CSF pleocytosis ^7^. Future outcome studies should consider these distinct disease patterns.

While plasma responses in viral meningitis had not been previously extensively evaluated, CSF cytokine profiles have been reported and are generally muted in PeV, and raised when pleocytosis is present ^26-28^. Low chemoattractant cytokine concentrations in PeV meningitis or EV without pleocytosis could be viewed as low levels of CNS inflammation, insufficient to recruit WBCs. Indeed, Harvala et al. proposed that many EV/PeV detections in CSF represent systemic infections with viral RNA leakage rather than true CNS infection ^18^. They reported higher CSF-to-plasma viral load ratios in infants without pleocytosis, driven mainly by plasma viral load differences. In light of our findings, we instead suggest that an aberrant immune response may fail to control plasma viral load, while muted CSF cytokine responses may reflect the contribution of WBCs to cytokine production in the CSF. Importantly, CSF cytokine responses are reduced but not absent in cases with an absence of pleocytosis and are still higher than in virus-negative controls ^26-28^. We conclude that RNA detection without pleocytosis is unlikely to be incidental and that reliance on pleocytosis as a diagnostic criterion for meningitis may overlook true CNS infection.

Another commonly proposed mechanism for the absence of CSF pleocytosis in infants is early presentation, before WBC migration into the CSF has occurred ^6,10,14^. While some studies support this association, they often include older children, introducing potential age-related bias ^13,14^. In our cohort, although fever duration was parent-reported and subject to recall bias, it was prospectively collected and clearly showed no association with CSF pleocytosis.

Furthermore, the plasma proteomic response was not influenced by time to presentation. Our findings argue against the hypothesis that early presentation accounts for the lack of pleocytosis in infant viral meningitis.

A key limitation of this study is its exploratory design. While we comprehensively characterised systemic immune and neuroinflammatory responses at presentation, no prespecified primary hypothesis or long-term outcomes were defined. Findings should therefore be interpreted as hypothesis-generating, and future studies with longitudinal follow-up are warranted to determine the clinical significance of these distinct immune response profiles. While the proteomic cohort was representative of the overall study population, inclusion depended on sample availability, introducing potential selection bias.

There is robust evidence that EV/PeV detection enables antibiotic discontinuation and decreases hospital stays, given the low IBI prevalence in viral meningitis infants ^1,2,4^. Our findings support this, with only 1/197 (0.5%) case of IBI in infants with virus-positive CSF. Blood PCR testing has been recommended to further identify this low-risk group, yet only 14% of febrile infants in this study underwent viral blood PCR, highlighting its limited use in UK clinical practice ^1,2,4^. The absence of routine EV/PeV blood PCR and lack of EV genotyping information are further limitations, as these prevent assessment of whether the EV immune heterogeneity reflects viral genotype differences or host-specific responses. Given the wide genetic diversity of EVs and known genotype-dependent differences in CSF/plasma viral loads, future work should also investigate the host response in patients with detected viremia along with the effects of EV genotype^14,17 18,29^.

Viral meningitis is a common cause of febrile illness in infants, yet its pathogenesis, particularly in cases without pleocytosis or neurological symptoms, remains unclear. Our findings suggest that absent CSF pleocytosis in EV and PeV meningitis is not due to early presentation or incidental viremia, but reflects a distinct immune phenotype marked by systemic cytokine activation and lymphocyte depletion. This response may limit leukocyte migration into the CSF despite systemic inflammation. These results challenge reliance on pleocytosis as a marker of CNS involvement, and we support a clinical definition for viral meningitis whereby the detection of EV or PeV in the CSF truly identifies a viral neurological disease, regardless of CSF pleocytosis. Future studies on outcomes to inform follow-up selection in infants with viral meningitis should explore these distinct immune responses.

## Contributors

CM, HG, EU and TW were involved in the conception and design of this study. EU, CM and TW coordinated the running of the original FIDO study, including data management and sample collection and storage. CM and HD undertook the analysis and verification of the analysis. CM wrote the first draft of the report with input from HG. All authors reviewed and approved the final manuscript and had full access to all the data in the study and had final responsibility for the decision to submit for publication.

## Data Availability

All data produced in the present study are available upon reasonable request to the authors

## Data sharing

De-identified participant data for this study will be made available to other investigators on reasonable request.

## Acknowledgments

We thank the funders of this study, Royal College of Emergency Medicine Doctoral Fellowship (RCEM 02/03/2021) and Sir Halley Stewart Trust (4292). We also thank all of the children and their families who participated in the FIDO study. We also thank all of the FIDO study hospital sites and staff who participated in screening and enrolment.

## Funding source

Royal College of Emergency Medicine Doctoral Fellowship (RCEM 02/03/2021) and Sir Halley Stewart Trust (4292). The funders played no part in the conception or design of this study

### AI Use Statement

Artificial intelligence (ChatGPT by OpenAI) was used to assist with improving the clarity, grammar, and readability of the manuscript. All AI-generated content was subsequently reviewed and edited by all authors to ensure scientific accuracy and integrity. No AI tools were used for data analysis or interpretation. The authors take full responsibility for the content of the published article.

