## Supplemental Figures and Tables for "Neuroinflammatory and immune responses in infants with enterovirus and parechovirus meningitis, and their association with CSF pleocytosis"

**Supplemental Figure 1. Study overview and analysis workflow.**

Flow diagram showing participant inclusion and analysis steps from the Febrile Infants Diagnostic Outcome (FIDO) cohort. EV = enterovirus; PeV = parechovirus; CSF = cerebrospinal fluid; LP = lumbar puncture; WBC = white blood cell; DEA = differential expression analysis; FC = fold change.

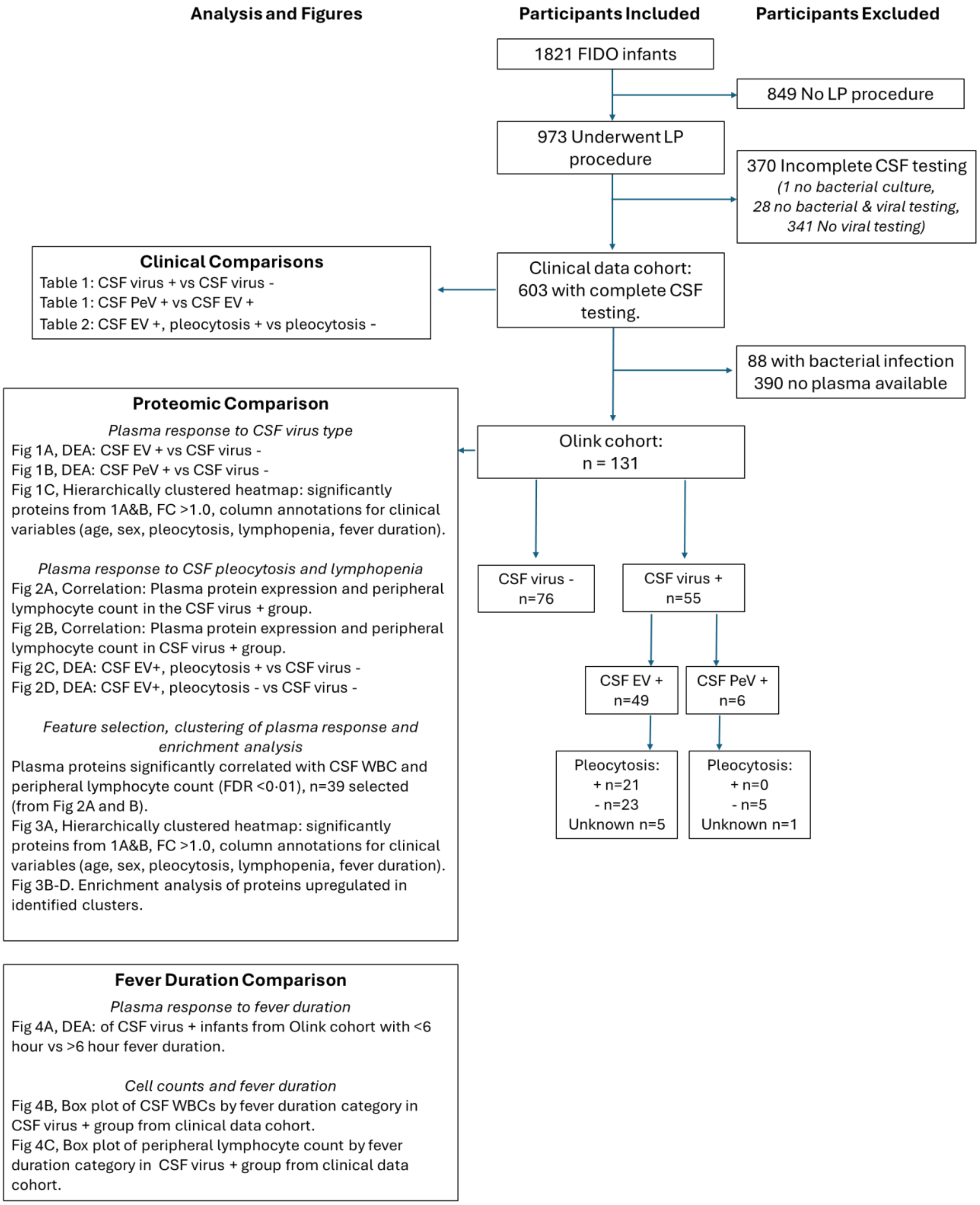

**Supplemental Figure 2. Box plots of significantly differentiated proteins in viral meningitis.** Box plots show scaled Olink plasma protein expression values across samples from infants with EV-positive, PeV-positive and virus-negative CSF. All proteins significantly differentiated between EV and virus-negative CSF groups, and the ten most significantly differentiated proteins between PeV and virus-negative CSF groups are shown. P-values from FDR adjusted differential expression analyses shown in Supplemental Table 2 and 3 are displayed as ns: adjusted p-value > 0.05; *: adjusted p-value <0.05; **: adjusted p-value < 0.01; ***: adjusted p-value ≤ 0.001.

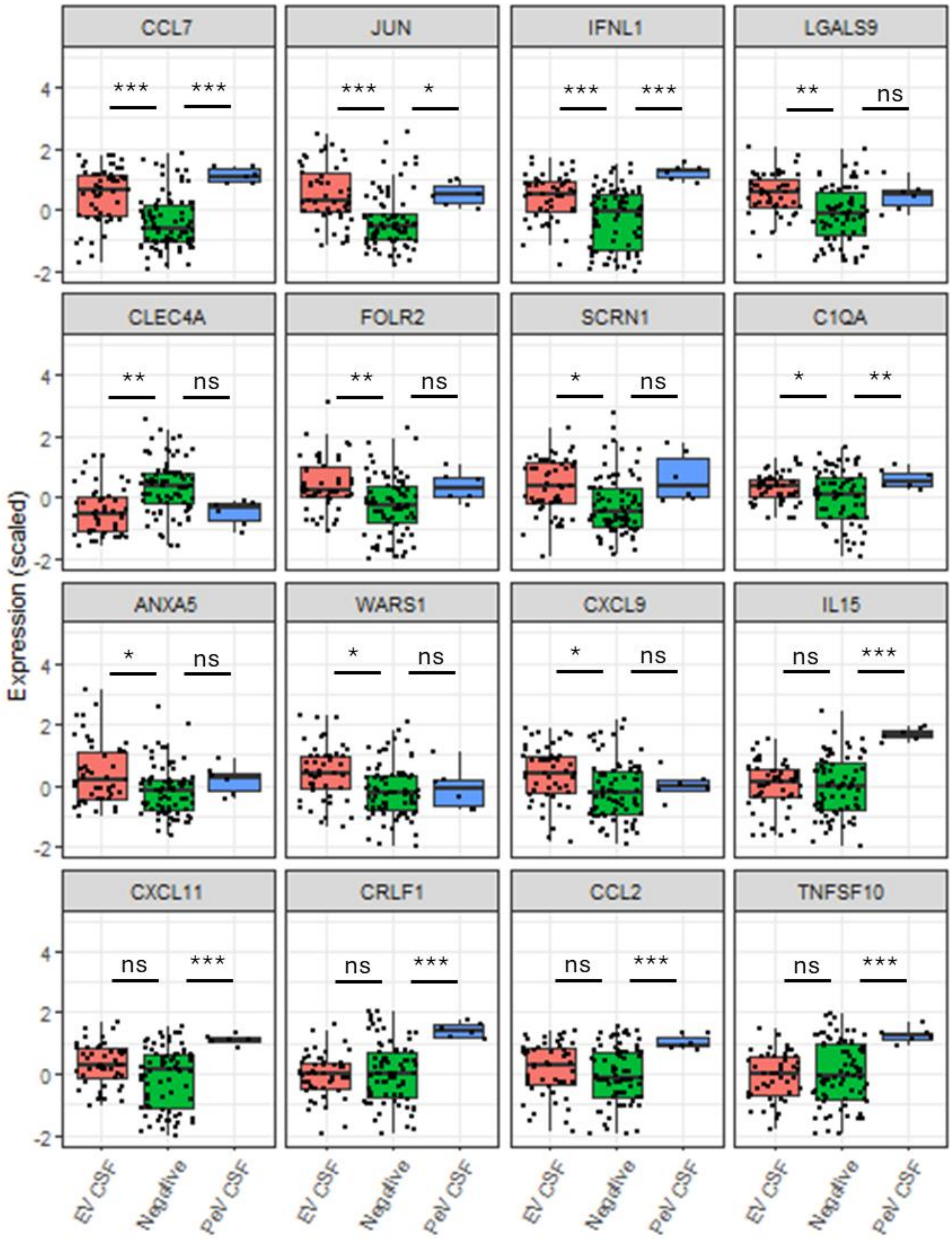

**Supplemental Figure 3. Scatter plots of plasma proteins significantly correlated with peripheral lymphocyte counts in viral meningitis.** Scatter plots show scaled Olink plasma protein expression values from infants with virus-positive CSF vs peripheral lymphocyte counts. The 8 most significantly positively and negatively correlated proteins are shown, from Supplemental Table 4.

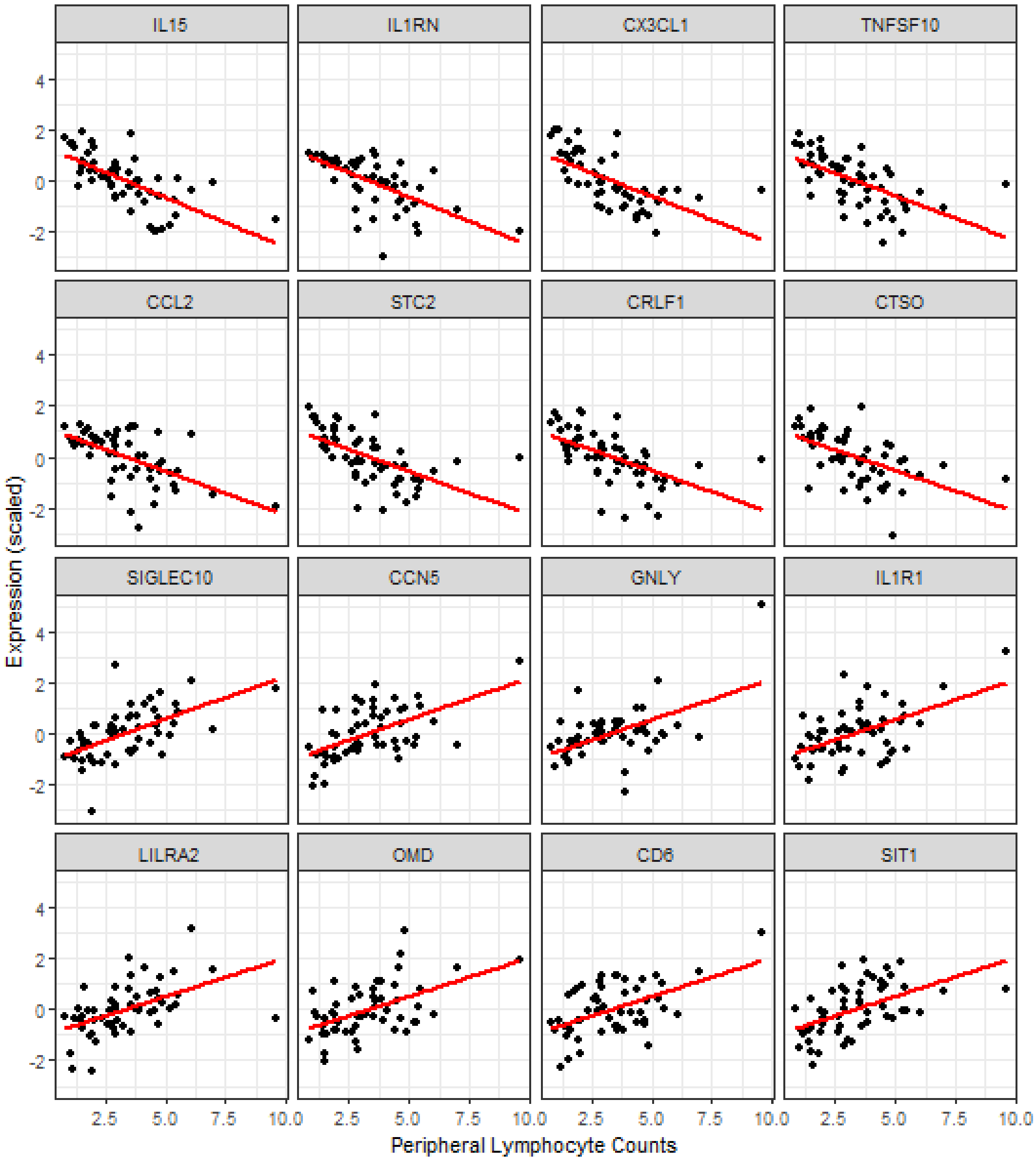

**Supplemental Figure 4. Scatter plots of plasma proteins significantly correlated with CSF WBC counts in viral meningitis.** Scatter plots show scaled Olink plasma protein expression values from infants with virus-positive CSF vs CSF WBC counts. The 8 most significantly negatively correlated proteins and the only three positively correlated proteins are shown, from Supplemental Table 5.

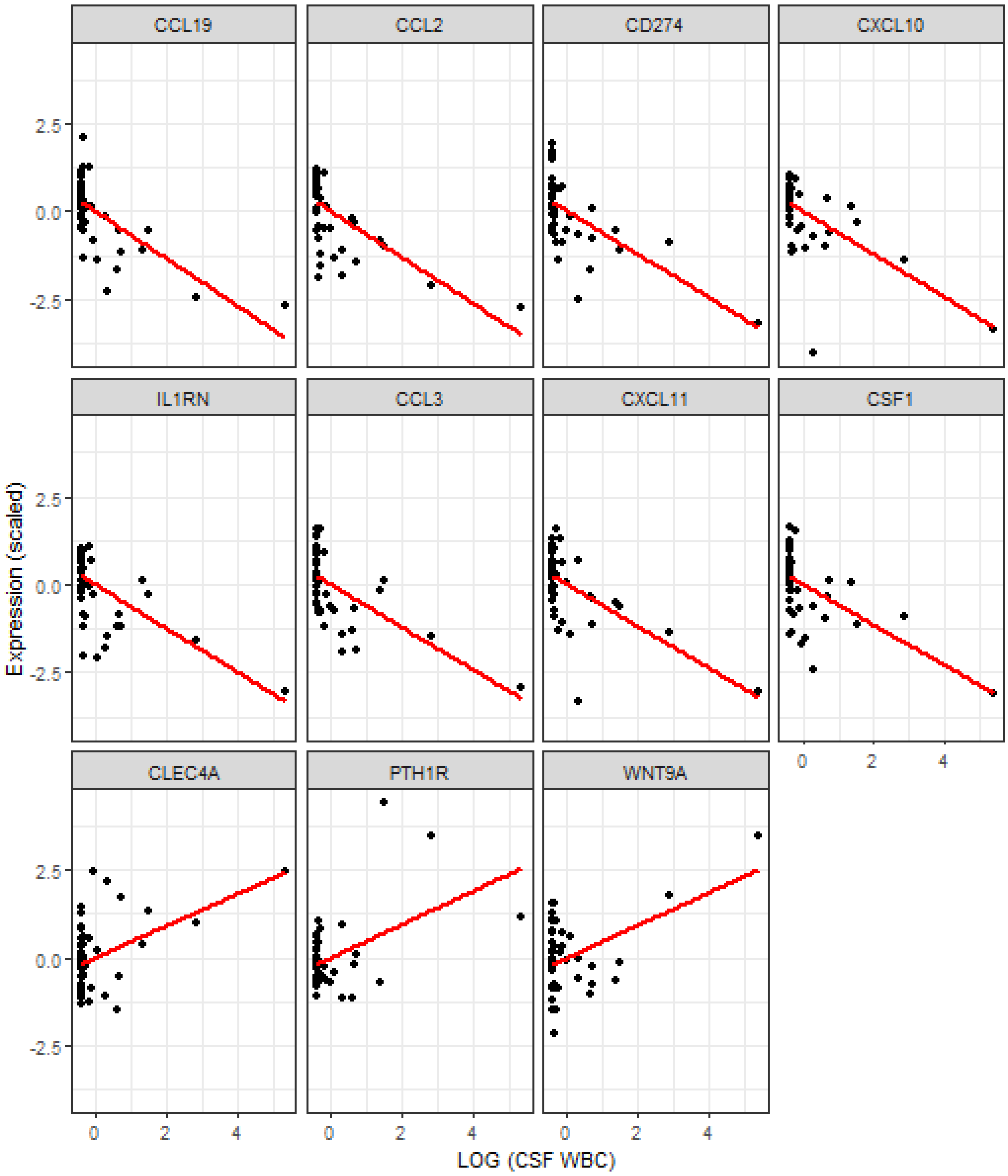

**Supplemental Figure 5. Box plots of significantly differentiated proteins in infants with EV meningitis with pleocytosis.** Box plots show scaled Olink plasma protein expression values across samples from infants with EV-positive CSF with and without pleocytosis, and virus-negative CSF. The ten most significantly upregulated proteins between EV with pleocytosis and virus-negative CSF groups are shown. Non-adjusted P-values from differential expression analyses shown in Supplemental Table 6 and 7 are displayed. ns: p-value > 0.05; *: p-value <0.05; **: p-value < 0.01; ***: p-value ≤ 0.001.

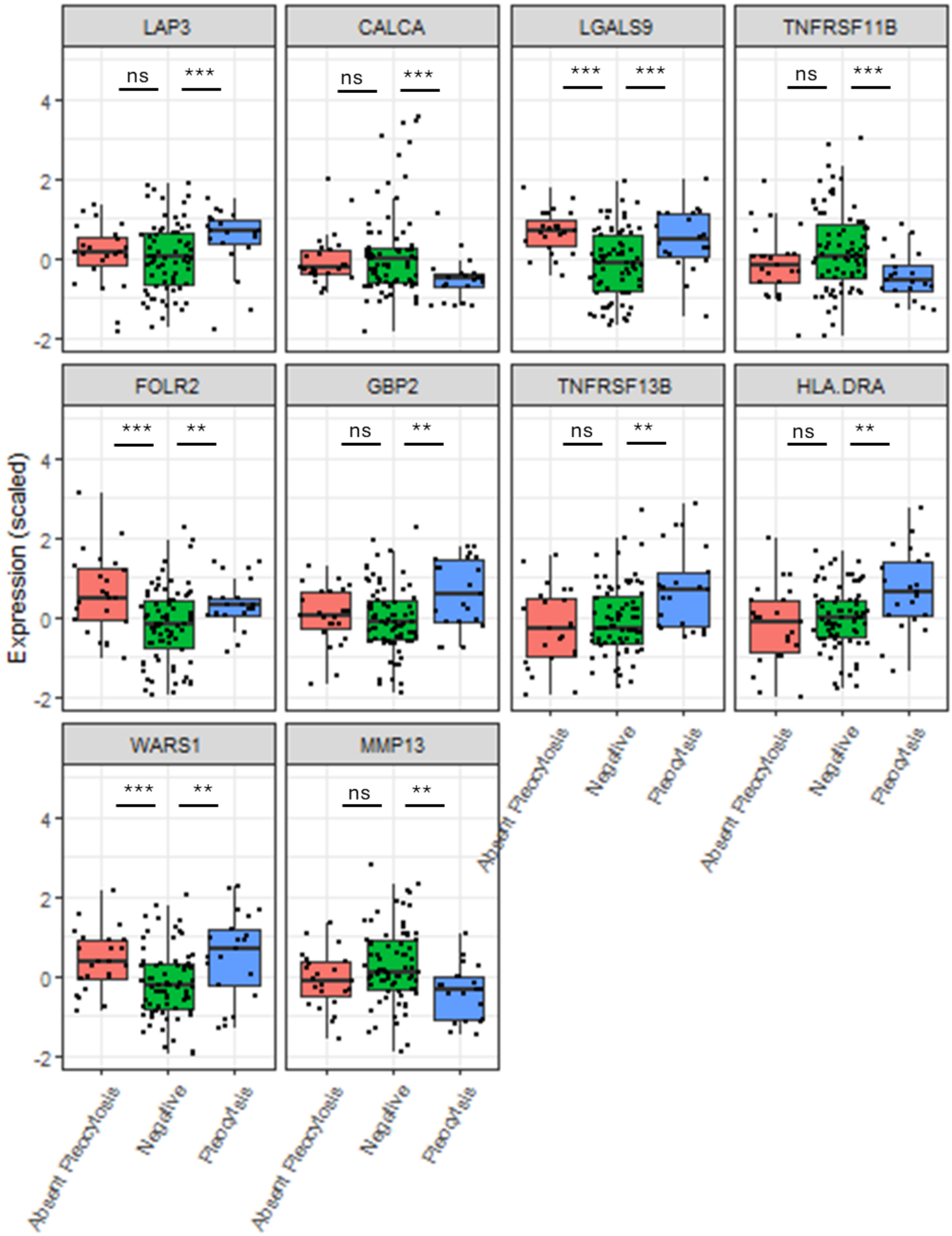

| | Olink cohort | Virus-positive CSF | Virus-negative CSF | P Value (Virus positive vs negative CSF) | PeV-positive CSF | EV-positive CSF | P Value (PeV vs EV CSF) |
|---|---|---|---|---|---|---|---|
| **Total**, n (%) | 131 | 55 (58) | 76 (41.9) | | 6 (10.9) | 49 (89.1) | |
| **Males**, n (%) | 80 (61) | 36 (65.5) | 44 (57.9) | 0.468 | 3 (50) | 80 (61) | 0.405 |
| **Age (days)** Median (IQR) | 41 (26-62) | 37 (24-56) | 49.5 (28.3-66.5) | 0.057 | 28.5 (23.8-36.5) | 39 (23.5-59.5) | 0.071 |
| **Temperature (°C)** Median (IQR) | 38.1 (37.7-38.5) | 38.2 (37.9-38.5) | 38.1 (37.6-38.5) | 0.042* | 38.2 (37.8-38.7) | 38.2 (37.8-38.5) | 0.520 |
| **CSF WBC count ($x10^9$/L)** Median (IQR) | n=120 1.0 (0.4-5.0) | n=49 4.9 (0.9-95.4) | n= 71 1.0 (0.3-3.0) | <0.001* | n=5 0.0 (0.0-2.1) | n=44 7.2 (1.0-128.0) | 0.007* |
| **Peripheral WBC count ($x10^9$/L)** Median (IQR) | n=127 8.2 (6.2-12.3) | n=54 8.2 (5.9-10.3) | n=73 8.6 (6.5-13.4) | 0.140 | n=6 6.1 (4.0-8.1) | n=48 8.3 (6.1-10.6) | 0.047* |
| **Peripheral lymphocyte count** ($x10^9$/L) Median (IQR) | n=131 3.1 (1.9-4.7) | n=54 3.0 (1.9-4.4) | n=73 3.7(2.2-5.6) | 0.214 | n=6 1.7 (1.0-2.4) | n=48 3.3 (2.1-4.6) | 0.009* |
| **CRP** (mg/L) Median (IQR) | n=131 7.0 (3.0-18.0) | n=55 8.0 (4.0-15.3) | n=76 5.5 (1.3-20.0) | 0.458 | n=6 4.0 (1.5-4.5) | n=49 10.0 (4.0-16.5) | 0.011* |

**Supplemental Table 1A. Patient characteristics and demographics of the Olink cohort.**

*Statistically significant. IQR, interquartile range; CSF, cerebrospinal fluid; WBC, white blood count; CRP, C-reactive protein; PeV, Parechovirus; EV Enterovirus.

| | Olink cohort | Remaining cohort without Olink analysis | Adjusted P value |
|---|---|---|---|
| **Total**, n (%) | 131 | 390 | |
| **Males**, n (%) | 80 (61) | 225 (57.7) | 0.260 |
| **Age (days)** Median (IQR) | 41 (26.0-62.0) | 38.0 (24.0–55.0) | 0.637 |
| **Temperature (°C)** Median (IQR) | 38.1 (37.7-38.5) | 38.1 (37.6–38.5) | 0.637 |
| **Unwell Appearance** n (%) | 90 (68.7) | 252 (64.6) | 0.325 |
| **Rash** n (%) | 36 (27.5) | 80 (20.6) | 0.454 |
| **Behaviour Change** n (%) | 107 (81.7) | 296 (76.1) | 0.959 |
| **Respiratory Symptoms**, n (%) | 51 (38.9) | 149 (38.2) | 0.637 |
| **Virus CSF,** n (%) | PeV: 6 (4.6)<br>EV: 49 (37.4) | PeV: 15 (3.8)<br>EV: 116 (29.8) | 0.568 |

**Supplemental Table 1B. Patient characteristics and demographic comparisons of the Olink cohort vs the reminder of the clinical data cohort which did not have samples available for Olink analysis.**

*Statistically significant. IQR, interquartile range; CSF, cerebrospinal fluid; PeV, Parechovirus; EV Enterovirus.

| Protein | UniProt ID | Panel | Fold Change | Adjusted p value |
|---|---|---|---|---|
| CCL7 | P80098 | Inflammation | 2.01 | <0.001 |
| JUN | P05412 | Inflammation | 1.01 | <0.001 |
| IFNL1 | Q8IU54 | Neurology | 1.89 | <0.001 |
| LGALS9 | O00182 | Inflammation | 0.43 | 0.001 |
| CLEC4A | Q9UMR7 | Inflammation | -0.33 | 0.003 |
| FOLR2 | P14207 | Neurology | 0.27 | 0.005 |
| SCRN1 | Q12765 | Inflammation | 0.65 | 0.019 |
| C1QA | P02745 | Inflammation | 0.20 | 0.019 |
| ANXA5 | P08758 | Neurology | 0.45 | 0.019 |
| WARS1 | P23381 | Neurology | 0.50 | 0.023 |
| CXCL9 | Q07325 | Inflammation | 0.71 | 0.030 |

**Supplemental Table 2**. Differentially expressed plasma proteins between infants with EV positive CSF vs virus negative CSF (FDR<0.05).

| Protein | UniProt ID | Panel | Fold Change | Adjusted p value |
|---|---|---|---|---|
| IL15 | P40933 | Inflammation | 1.53 | <0.001 |
| CXCL11 | O14625 | Neurology | 2.92 | <0.001 |
| IL1RN | P18510 | Inflammation | 1.68 | <0.001 |
| CXCL10 | P02778 | Inflammation | 2.59 | <0.001 |
| CX3CL1 | P78423 | Neurology | 1.32 | <0.001 |
| IFNL1 | Q8IU54 | Neurology | 3.46 | <0.001 |
| CCL7 | P80098 | Inflammation | 3.37 | <0.001 |
| CRLF1 | O75462 | Inflammation | 0.55 | <0.001 |
| CCL2 | P13500 | Neurology | 1.50 | <0.001 |
| TNFSF10 | P50591 | Inflammation | 1.11 | <0.001 |
| SIGLEC10 | Q96LC7 | Inflammation | -0.59 | <0.001 |
| TNFRSF10B | O14763 | Neurology | -0.62 | <0.001 |
| CD274 | Q9NZQ7 | Neurology | 1.37 | <0.001 |
| CSF1 | P09603 | Inflammation | 0.46 | <0.001 |
| CTSO | P43234 | Inflammation | 1.08 | 0.001 |
| TNFSF11 | O14788 | Inflammation | 1.26 | 0.002 |
| STC2 | O76061 | Neurology | 0.55 | 0.003 |
| CD6 | P30203 | Inflammation | -1.00 | 0.003 |
| SCARB2 | Q14108 | Neurology | 0.73 | 0.003 |
| LY9 | Q9HBG7 | Inflammation | -0.38 | 0.004 |
| C1QA | P02745 | Inflammation | 0.30 | 0.008 |
| HSD11B1 | P28845 | Inflammation | -0.66 | 0.009 |
| COPE | O14579 | Neurology | -0.60 | 0.016 |
| CCL23 | P55773 | Inflammation | -0.76 | 0.025 |
| EREG | O14944 | Neurology | -0.74 | 0.025 |
| JUN | P05412 | Inflammation | 0.91 | 0.026 |
| PVR | P15151 | Neurology | 0.27 | 0.026 |
| CCL24 | O00175 | Inflammation | -1.31 | 0.026 |
| KLRD1 | Q13241 | Inflammation | -0.36 | 0.031 |

| | | | | |
|---|---|---|---|---|
| LAMP3 | Q9UQV4 | Inflammation | 0.60 | 0.034 |
| IL1R1 | P14778 | Neurology | -0.41 | 0.037 |
| IL22RA1 | Q8N6P7 | Inflammation | 0.77 | 0.043 |

**Supplemental Table 3.** Differentially expressed plasma proteins between infants with PeV positive CSF vs virus negative CSF (FDR<0.05).

| Protein | UniProt ID | Panel | Pearsons R | Adjusted p value |
|---|---|---|---|---|
| IL15 | P78423 | Neurology | -0.668 | <0.001 |
| IL1RN | P50591 | Inflammation | -0.644 | <0.001 |
| CX3CL1 | P13500 | Neurology | -0.622 | <0.001 |
| TNFSF10 | O76061 | Neurology | -0.598 | <0.001 |
| CCL2 | Q96LC7 | Inflammation | -0.577 | <0.001 |
| STC2 | O75462 | Inflammation | -0.574 | <0.001 |
| SIGLEC10 | O76076 | Neurology | 0.571 | <0.001 |
| CRLF1 | P43234 | Inflammation | -0.557 | 0.001 |
| CCN5 | P22749 | Neurology | 0.550 | 0.001 |
| CTSO | P14778 | Neurology | -0.544 | 0.002 |
| GNLY | Q99538 | Inflammation | 0.539 | 0.002 |
| IL1R1 | Q8N149 | Neurology | 0.532 | 0.002 |
| LGMN | Q99983 | Inflammation | -0.512 | 0.003 |
| LILRA2 | P30203 | Inflammation | 0.512 | 0.003 |
| OMD | Q9Y3P8 | Inflammation | 0.512 | 0.003 |
| CD6 | P56279 | Neurology | 0.512 | 0.003 |
| SIT1 | P24387 | Inflammation | 0.515 | 0.003 |
| TCL1A | P05231 | Inflammation | 0.516 | 0.003 |
| CRHBP | O14836 | Inflammation | 0.519 | 0.003 |
| IL6 | O60880 | Inflammation | -0.504 | 0.004 |
| TNFRSF13B | P23582 | Inflammation | 0.504 | 0.004 |
| SH2D1A | Q9NZQ7 | Neurology | 0.489 | 0.006 |
| NPPC | O14625 | Neurology | -0.488 | 0.006 |
| CD274 | Q8IU54 | Neurology | -0.482 | 0.007 |
| CXCL11 | P80098 | Inflammation | -0.482 | 0.007 |
| IFNL1 | P13236 | Inflammation | -0.479 | 0.007 |
| CCL7 | O14763 | Neurology | -0.470 | 0.009 |
| CCL4 | P24001 | Inflammation | -0.469 | 0.009 |
| TNFRSF10B | P02778 | Inflammation | 0.466 | 0.009 |
| IL32 | Q14116 | Inflammation | 0.462 | 0.011 |
| CXCL10 | P02778 | Inflammation | -0.458 | 0.011 |
| IL18 | P33241 | Inflammation | 0.458 | 0.011 |
| HLA.E | Q86WV1 | Neurology | -0.452 | 0.013 |
| LSP1 | Q6DN72 | Inflammation | 0.436 | 0.021 |
| SKAP1 | P25774 | Neurology | 0.433 | 0.022 |
| FCRL6 | Q9H5V8 | Neurology | 0.433 | 0.022 |
| CTSS | Q92752 | Neurology | -0.432 | 0.022 |
| CDCP1 | P10147 | Inflammation | 0.426 | 0.025 |

| | | | | |
|---|---|---|---|---|
| TNR | Q9Y6Q6 | Inflammation | 0.421 | 0.028 |
| CCL3 | Q9BXJ7 | Inflammation | -0.419 | 0.029 |
| TNFRSF11A | P51671 | Inflammation | -0.414 | 0.032 |
| AMN | P09341 | Inflammation | -0.414 | 0.032 |
| CCL11 | P13647 | Neurology | -0.414 | 0.032 |
| CXCL1 | Q5JZY3 | Neurology | -0.411 | 0.033 |
| KRT5 | P01258 | Neurology | 0.410 | 0.033 |
| EPHA10 | Q9BZW8 | Inflammation | 0.408 | 0.034 |
| CALCA | Q14005 | Inflammation | -0.407 | 0.035 |
| CD244 | P28799 | Neurology | 0.401 | 0.039 |
| IL16 | Q6NW40 | Neurology | 0.402 | 0.039 |
| GRN | P28908 | Neurology | -0.396 | 0.044 |
| RGMB | Q15109 | Inflammation | -0.393 | 0.045 |
| TNFRSF8 | O95727 | Neurology | 0.394 | 0.045 |
| AGER | P19438 | Neurology | -0.390 | 0.048 |
| CRTAM | Q9UHC6 | Inflammation | 0.390 | 0.048 |
| TNFRSF1A | P01732 | Neurology | -0.388 | 0.048 |
| CNTNAP2 | P12532 | Inflammation | -0.387 | 0.048 |
| CD8A | P18564 | Inflammation | 0.387 | 0.048 |
| CKMT1A_CKMT1B | O00468 | Inflammation | 0.388 | 0.048 |

**Supplemental Table 4.** Plasma proteins significantly correlated with peripheral lymphocyte counts (FDR<0.05).

| Protein | UniProt ID | Panel | Pearsons R | Adjusted p value |
|---|---|---|---|---|
| CCL19 | Q99731 | Neurology | -0.67 | <0.001 |
| CCL2 | P13500 | Neurology | -0.65 | <0.001 |
| CD274 | Q9NZQ7 | Neurology | -0.61 | <0.001 |
| CXCL10 | P02778 | Inflammation | -0.61 | <0.001 |
| IL1RN | P18510 | Inflammation | -0.62 | <0.001 |
| CCL3 | P10147 | Inflammation | -0.60 | <0.001 |
| CXCL11 | O14625 | Neurology | -0.59 | <0.001 |
| CSF1 | P09603 | Inflammation | -0.57 | 0.00 |
| CCL11 | P51671 | Inflammation | -0.56 | 0.00 |
| TNFRSF6B | O95407 | Neurology | -0.55 | 0.00 |
| CCL20 | P78556 | Inflammation | -0.54 | 0.00 |
| CRLF1 | O75462 | Inflammation | -0.54 | 0.00 |
| STC2 | O76061 | Neurology | -0.53 | 0.00 |
| CCL4 | P13236 | Inflammation | -0.52 | 0.01 |
| IFNG | P01579 | Inflammation | -0.52 | 0.01 |
| IFNL1 | Q8IU54 | Neurology | -0.52 | 0.01 |
| TNFSF10 | P50591 | Inflammation | -0.52 | 0.01 |
| TNF | P01375 | Neurology | -0.51 | 0.01 |
| CCL7 | P80098 | Inflammation | -0.48 | 0.02 |
| CXCL9 | Q07325 | Inflammation | -0.48 | 0.02 |
| PTH1R | Q03431 | Inflammation | 0.48 | 0.02 |

| | | | | |
|---|---|---|---|---|
| GRN | P28799 | Neurology | -0.47 | 0.02 |
| WNT9A | O14904 | Inflammation | 0.47 | 0.02 |
| CTSS | P25774 | Neurology | -0.46 | 0.02 |
| BTN2A1 | Q7KYR7 | Inflammation | -0.46 | 0.03 |
| CLEC4A | Q9UMR7 | Inflammation | 0.45 | 0.03 |
| SCRN1 | Q12765 | Inflammation | -0.45 | 0.03 |
| CLEC4C | Q8WTT0 | Inflammation | -0.45 | 0.03 |
| CXCL1 | P09341 | Inflammation | -0.45 | 0.03 |
| TNFRSF1A | P19438 | Neurology | -0.45 | 0.03 |
| CXCL6 | P80162 | Inflammation | -0.44 | 0.03 |
| TPPP3 | Q9BW30 | Neurology | -0.44 | 0.04 |
| CD74 | P04233 | Neurology | -0.43 | 0.04 |

**Supplemental Table 5.** Plasma proteins significantly correlated with CSF WBC counts (FDR<0.05).

| Protein | UniProt ID | Panel | Fold Change | Adjusted p value |
|---|---|---|---|---|
| CCL7 | P80098 | Inflammation | 3.058 | <0.001 |
| IFNL1 | Q8IU54 | Neurology | 2.542 | <0.001 |
| CXCL10 | P02778 | Inflammation | 2.114 | <0.001 |
| CCL2 | P13500 | Neurology | 1.215 | <0.001 |
| CXCL9 | Q07325 | Inflammation | 1.268 | <0.001 |
| JUN | P05412 | Inflammation | 1.424 | <0.001 |
| LGALS9 | O00182 | Inflammation | 0.545 | <0.001 |
| CXCL11 | O14625 | Neurology | 1.797 | <0.001 |
| CLEC4A | Q9UMR7 | Inflammation | -0.427 | <0.001 |
| IL1RN | P18510 | Inflammation | 1.182 | <0.001 |
| IFNG | P01579 | Inflammation | 2.211 | <0.001 |
| IL17C | Q9P0M4 | Inflammation | -0.874 | <0.001 |
| SCRN1 | Q12765 | Inflammation | 0.961 | <0.001 |
| IL33 | O95760 | Inflammation | 0.754 | 0.001 |
| FOSB | P53539 | Neurology | 0.648 | 0.001 |
| SIGLEC5 | O15389 | Neurology | 1.136 | 0.001 |
| IL1R1 | P14778 | Neurology | -0.296 | 0.002 |
| TPPP3 | Q9BW30 | Neurology | 0.984 | 0.002 |
| TNFRSF6B | O95407 | Neurology | 0.501 | 0.002 |
| SIGLEC1 | Q9BZZ2 | Inflammation | 0.507 | 0.002 |
| CSF1 | P09603 | Inflammation | 0.272 | 0.003 |
| C1QA | P02745 | Inflammation | 0.233 | 0.009 |
| PAPPA | Q13219 | Inflammation | -0.370 | 0.017 |
| CD274 | Q9NZQ7 | Neurology | 0.474 | 0.017 |
| GRN | P28799 | Neurology | 0.380 | 0.021 |
| LILRB4 | Q8NHJ6 | Inflammation | 0.435 | 0.022 |
| WARS1 | P23381 | Neurology | 0.526 | 0.024 |
| FOLR2 | P14207 | Neurology | 0.320 | 0.024 |
| IL15 | P40933 | Inflammation | 0.468 | 0.025 |
| AGRN | O00468 | Inflammation | 0.396 | 0.026 |

| | | | | |
|---|---|---|---|---|
| CCL19 | Q99731 | Neurology | 0.487 | 0.028 |
| LAIR1 | Q6GTX8 | Inflammation | 0.352 | 0.033 |
| SCARB2 | Q14108 | Neurology | 0.378 | 0.034 |
| SIT1 | Q9Y3P8 | Inflammation | -0.606 | 0.036 |
| ANXA5 | P08758 | Neurology | 0.575 | 0.044 |

**Supplemental Table 6.** Differentially expressed plasma proteins between infants with EV positive CSF without pleocytosis vs virus negative CSF (FDR<0.05).

| Protein | UniProt ID | Panel | Fold Change | p value | Adjusted p value |
|---|---|---|---|---|---|
| LAP3 | P28838 | Inflammation | 0.59 | <0.001 | 0.137 |
| CALCA | P01258 | Neurology | -0.69 | <0.001 | 0.137 |
| LGALS9 | O00182 | Inflammation | 0.43 | 0.001 | 0.137 |
| TNFRSF11B | O00300 | Inflammation | -0.37 | 0.001 | 0.137 |
| FOLR2 | P14207 | Neurology | 0.24 | 0.001 | 0.137 |
| GBP2 | P32456 | Inflammation | 0.98 | 0.001 | 0.137 |
| TNFRSF13B | O14836 | Inflammation | 0.43 | 0.002 | 0.159 |
| HLA-DRA | P01903 | Inflammation | 0.66 | 0.002 | 0.159 |
| WARS1 | P23381 | Neurology | 0.68 | 0.002 | 0.159 |
| MMP13 | P45452 | Neurology | -0.55 | 0.002 | 0.159 |
| C1QA | P02745 | Inflammation | 0.18 | 0.002 | 0.159 |
| SOD2 | P04179 | Neurology | 0.72 | 0.004 | 0.224 |
| JUN | P05412 | Inflammation | 0.62 | 0.004 | 0.224 |
| IFNL1 | Q8IU54 | Neurology | 1.24 | 0.004 | 0.224 |
| FSTL3 | O95633 | Inflammation | -0.35 | 0.005 | 0.229 |
| ADAM23 | O75077 | Inflammation | -0.27 | 0.006 | 0.229 |
| CCL7 | P80098 | Inflammation | 1.41 | 0.006 | 0.229 |
| NUDC | Q9Y266 | Inflammation | 0.36 | 0.006 | 0.229 |
| IL17RB | Q9NRM6 | Inflammation | 0.44 | 0.007 | 0.229 |
| CDH15 | P55291 | Neurology | -0.39 | 0.007 | 0.229 |
| STC2 | O76061 | Neurology | -0.26 | 0.007 | 0.229 |
| ANXA5 | P08758 | Neurology | 0.41 | 0.007 | 0.229 |
| CLEC4A | Q9UMR7 | Inflammation | -0.27 | 0.008 | 0.229 |
| IL2 | P60568 | Inflammation | -0.41 | 0.008 | 0.229 |
| IL24 | Q13007 | Inflammation | -0.29 | 0.008 | 0.229 |
| CLEC4C | Q8WTT0 | Inflammation | -0.41 | 0.008 | 0.229 |
| TNFSF10 | P50591 | Inflammation | -0.48 | 0.009 | 0.231 |

**Supplemental Table 7** Differentially expressed plasma proteins between infants with EV-positive CSF with pleocytosis vs virus-negative CSF P<0.01, without FDR applied.

| Protein | UniProt ID | FC 2 vs 1 | p value 2 vs 1 | FC 3 vs 1 | p value 3 vs 1 | FC 2 vs 3 | p value 2 vs 3 | Upregulated protein | | |
|---|---|---|---|---|---|---|---|---|---|---|
| | | | | | | | | Cluster 1 | Cluster 2 | Cluster 3 |
| CCL11 | P51671 | 0.80 | <0.001 | 0.72 | <0.001 | 0.08 | 0.646 | No | Yes | Yes |
| CCL19 | Q99731 | 1.25 | <0.001 | 1.31 | <0.001 | -0.06 | 0.729 | No | Yes | Yes |
| CCL2 | P13500 | 2.18 | <0.001 | 1.93 | <0.001 | 0.25 | 0.146 | No | Yes | Yes |
| CCL3 | P10147 | 0.86 | <0.001 | 0.85 | <0.001 | 0.01 | 0.970 | No | Yes | Yes |
| CCL4 | P13236 | 0.78 | <0.001 | 0.74 | <0.001 | 0.04 | 0.806 | No | Yes | Yes |
| CCL7 | P80098 | 2.83 | <0.001 | 2.82 | <0.001 | 0.01 | 0.980 | No | Yes | Yes |
| CD274 | Q9NZQ7 | 1.28 | <0.001 | 0.87 | <0.001 | 0.42 | 0.010 | No | Yes | Yes |
| CRLF1 | O75462 | 0.63 | <0.001 | 0.38 | <0.001 | 0.26 | 0.001 | No | Yes | Yes |
| CSF1 | P09603 | 0.44 | <0.001 | 0.43 | <0.001 | 0.02 | 0.832 | No | Yes | Yes |
| CTSO | P43234 | 1.18 | <0.001 | 0.73 | <0.001 | 0.44 | 0.009 | No | Yes | Yes |
| CX3CL1 | P78423 | 1.35 | <0.001 | 0.61 | <0.001 | 0.73 | <0.001 | No | Yes | Yes |
| CXCL10 | P02778 | 2.60 | <0.001 | 2.17 | <0.001 | 0.43 | 0.006 | No | Yes | Yes |
| CXCL11 | O14625 | 2.88 | <0.001 | 2.42 | <0.001 | 0.45 | 0.186 | No | Yes | Yes |
| CXCL9 | Q07325 | 1.13 | <0.001 | 1.10 | <0.001 | 0.04 | 0.892 | No | Yes | Yes |
| IFNG | P01579 | 1.64 | 0.008 | 1.95 | 0.002 | -0.30 | 0.620 | No | Yes | Yes |
| IFNL1 | Q8IU54 | 2.59 | <0.001 | 1.91 | <0.001 | 0.68 | 0.047 | No | Yes | Yes |
| IL15 | P40933 | 1.33 | <0.001 | 0.78 | <0.001 | 0.55 | 0.004 | No | Yes | Yes |
| IL1RN | P18510 | 2.39 | <0.001 | 1.80 | <0.001 | 0.59 | <0.001 | No | Yes | Yes |
| IL6 | P05231 | 1.97 | <0.001 | 1.92 | <0.001 | 0.04 | 0.930 | No | Yes | Yes |
| LGMN | Q99538 | 1.00 | <0.001 | 0.59 | <0.001 | 0.41 | 0.012 | No | Yes | Yes |
| STC2 | O76061 | 0.75 | <0.001 | 0.41 | <0.001 | 0.33 | <0.001 | No | Yes | Yes |
| TNF | P01375 | 0.48 | 0.003 | 0.71 | <0.001 | -0.23 | 0.151 | No | Yes | Yes |
| TNFRSF6B | O95407 | 0.79 | <0.001 | 1.16 | <0.001 | -0.37 | 0.028 | No | Yes | Yes |
| TNFSF10 | P50591 | 1.42 | <0.001 | 0.95 | <0.001 | 0.47 | 0.005 | No | Yes | Yes |
| CCN5 | O76076 | -0.48 | 0.001 | 0.04 | 0.786 | -0.52 | <0.001 | Yes | No | Yes |
| CD6 | P30203 | -0.67 | 0.004 | -0.18 | 0.440 | -0.49 | 0.024 | Yes | No | Yes |
| CRHBP | P24387 | -0.53 | <0.001 | -0.07 | 0.533 | -0.45 | 0.003 | Yes | No | Yes |
| LILRA2 | Q8N149 | -0.49 | <0.001 | 0.15 | 0.325 | -0.64 | <0.001 | Yes | No | Yes |
| OMD | Q99983 | -0.07 | 0.020 | 0.04 | 0.285 | -0.10 | 0.003 | Yes | No | Yes |
| SH2D1A | O60880 | -0.88 | <0.001 | -0.17 | 0.492 | -0.70 | 0.005 | Yes | No | Yes |
| SIGLEC10 | Q96LC7 | -0.55 | <0.001 | 0.03 | 0.813 | -0.58 | <0.001 | Yes | No | Yes |
| SIT1 | Q9Y3P8 | -0.85 | <0.001 | 0.01 | 0.963 | -0.86 | <0.001 | Yes | No | Yes |
| TCL1A | P56279 | -1.23 | <0.001 | 0.16 | 0.500 | -1.38 | <0.001 | Yes | No | Yes |
| TNFRSF10B | O14763 | -0.43 | <0.001 | -0.19 | 0.076 | -0.24 | 0.039 | Yes | No | Yes |
| TNFRSF13B | O14836 | -0.74 | <0.001 | -0.26 | 0.110 | -0.49 | <0.001 | Yes | No | Yes |
| CCL20 | P78556 | 0.51 | 0.111 | 1.11 | 0.006 | -0.59 | 0.042 | No | No | Yes |
| NPPC | P23582 | 0.57 | <0.001 | 0.11 | 0.437 | 0.47 | <0.001 | No | Yes | No |
| GNLY | P22749 | -0.79 | 0.048 | -0.45 | 0.314 | -0.34 | 0.170 | Yes | No | No |
| IL1R1 | P14778 | -0.43 | <0.001 | -0.28 | 0.036 | -0.16 | 0.087 | Yes | No | Yes |

**Supplemental Table 8.** Differentially expressed plasma proteins between infants in cluster 1, 2 and 3 from Figure 3B. Proteins are marked as upregulated in a cluster if they are significantly (P<0.05) upregulated in any cluster comparison.
